# Genetic Contributions to Alzheimer’s Disease and Frontotemporal Dementia in Admixed Latin American Populations

**DOI:** 10.1101/2024.10.29.24315197

**Authors:** Juliana Acosta-Uribe, Stefanie D. Piña Escudero, J. Nicholas Cochran, Jared W. Taylor, P. Alejandra Castruita, Caroline Jonson, Erin A. Barinaga, Kevin Roberts, Alexandra R. Levine, Dawwod S. George, José Alberto ÁvilaFunes, María I. Behrens, Martín A. Bruno, Luis I. Brusco, Nilton Custodio, Claudia Duran-Aniotz, Francisco Lopera, Diana L. Matallana, Andrea Slachevsky, Leonel T. Takada, Lina M. Zapata-Restrepo, Dafne E. Durón-Reyes, Elisa de Paula França Resende, Nancy Gelvez, Maria E. Godoy, Marcelo A. Maito, Shireen Javandel, Bruce L. Miller, Mike A. Nalls, Hampton Leonard, Dan Vitale, Sara Bandres-Ciga, Mathew J. Koretsky, Andrew B. Singleton, Caroline B. Pantazis, Victor Valcour, Agustin Ibañez, Kenneth S. Kosik, Jennifer S. Yokoyama, the Multi-Partner Consortium to Expand Dementia Research in Latin America (ReDLat)

**Author notes:** Both authors contributed equally (Juliana Acosta-Uribe, Stefanie D. Piña Escudero). Passed away during the preparation of this manuscript on September 10, 2024. Both authors contributed equally (Kenneth S. Kosik, Jennifer S. Yokoyama). **Corresponding author** Jennifer S. Yokoyama Address for correspondence: 1651 4th St. Room 3C08, San Francisco, CA 94158. USA.

## Abstract

**Background:** Latin America’s diverse genetic makeup, shaped by centuries of admixture, presents a unique opportunity to study Alzheimer’s disease dementia (AD) and frontotemporal dementia (FTD). Our aim is to identify genetic variations associated with AD and FTD within this population.

**Methods:** The Multi-Partner Consortium to Expand Dementia Research in Latin America (ReDLat) recruited 2,162 participants with AD, FTD, and healthy controls from six Latin American countries (Argentina, Brazil, Chile, Colombia, Mexico, and Peru). All participants underwent array, exome, and/or whole-genome sequencing. Population structure was analyzed using Principal Component Analysis and ADMIXTURE, projecting the ReDLat population onto the 1000 Genomes Project database. To identify genes associated with autosomal dominant, autosomal recessive, or X-linked forms of adult-onset dementia, we searched the Online Mendelian Inheritance in Man database and analyzed pedigree information. Variant interpretation followed guidelines from the American College of Medical Genetics and Genomics, and the Guerreiro algorithm was applied for the *PSEN1* and *PSEN2* genes.

**Results:** Global ancestry analysis of the ReDLat cohort revealed a predominant mix of American, African, and European ancestries. Uniquely, Brazil displayed an additional East Asian component accurately reflecting the historical admixture patterns from this region. We identified 17 pathogenic variants, a pathogenic *C9orf72* expansion, and 44 variants of uncertain significance. Among our cohort, 70 families exhibited autosomal dominant inheritance of neurodegenerative diseases, with 48 families affected by AD and 22 by FTD. In families with AD, We discovered a novel variant in the *PSEN1* gene, c.519G>T (p.Leu173Phe), along with other previously described variants seen in the region, such as c.356C>T (p.Thr119Ile). In families with FTD, the most commonly associated gene was *GRN*, followed by *MAPT*. Notably, we identified a patient meeting criteria for FTD who carried a pathogenic variant in *SOD1*, c.388G>A (p.Phe21Leu), which had previously been reported in another FTD patient from the same geographical region.

**Conclusions:** This study provides the first snapshot of genetic contributors to AD and FTD in a multisite cohort across Latin America. It will be critical to evaluate the generalizability of genetic risk factors for AD and FTD across diverse ancestral backgrounds, considering distinct social determinants of health and accounting for modifiable risk factors that may influence disease risk and resilience across different cultures.

## Background

Latin America houses a unique blend of Indigenous American, European, and African ancestries, creating a diverse genetic landscape that emerged from the conquest of the Americas and the slave trade that began in the 1500s. (1) Significant East Asian immigration that occurred during the early 20th century has further contributed to the continent’s population diversity as observed today. (2,3) This admixture harbors a spectrum of novel genetic variants, including some that may modulate susceptibility to neurodegenerative diseases like Alzheimer’s disease (AD) and frontotemporal dementia (FTD), or may result in a higher allelic frequency of known risk-conveying variants for neurodegeneration. (4) Studying these populations, with their complex genetic architecture, offers an invaluable resource for understanding neurodegenerative diseases.

Assessing admixed populations is particularly interesting because admixture introduces a rich heterogeneity of alleles, which can be crucial for understanding genetic risk. For instance, in the Colombian population, researchers from a single center in Medellín have identified 13 different pathogenic *PSEN1* variants. (5) This contrasts with the nine independent variants described in a screening study from nine centers throughout the Iberian Peninsula. (6) It is hypothesized that these mutations, which arose in different ancestral backgrounds, led to high amyloid-beta levels which possibly conferred antimicrobial benefits to carriers during the American Conquest. (5) Additionally, in the Peruvian population, a recent genome-wide association study and functional analysis suggested that the *NFASC* gene, located on chromosome 1, is associated with AD. The *NFASC* locus showed significant contributions from both European and African ancestries. (7) This finding emphasizes the crucial role that diverse ancestral backgrounds play in both uncovering novel variation and understanding the complex genetic foundations of AD.

Due to cultural, religious, and historical factors, Latin American families have traditionally been large. Government policies during the 1960s and 1970s further encouraged population growth, resulting in an average family size of six children. (8,9) Additionally, geographic and socioeconomic conditions often led to extended families residing in close proximity. (8,10) This unique demographic structure provides an exceptional opportunity to study large families, tracing the lineage and impact of specific genetic variants across generations. For example, the *PSEN1* E280A (Glu280Ala) variant in Medellín, Colombia has been traced back to approximately 500 years ago, around the time of the Spanish invasion. This variant entered an admixed population with a small effective population size, generating a large founder effect. This historical and genetic tracing aids in understanding the genetic drift that occurs over time due to the small population size and increases the probability of identifying both risk and protective genetic variants. (11–13)

The scientific community from the region recognized the immense potential of this approach and formed the Multi-Partner Consortium to Expand Dementia Research in Latin America (ReDLat). (14) This consortium fosters collaboration among researchers, clinicians, and institutions across six Latin American countries and the United States to leverage the unique genetic diversity and demographic characteristics that influence AD and FTD in Latin American populations. ReDLat aims to highlight the unique genetic diversity and demographic features of these populations associated with these neurodegenerative diseases. (15) Ultimately, the consortium’s work seeks to refine diagnostic approaches, to develop targeted therapeutic interventions, and significantly enhance our understanding of dementia across Latin America. (14)

This paper is the first genetic report from this consortium, focusing on AD and FTD in admixed Latin American participants, with a particular emphasis on families. Our research efforts aim to identify genetic variants associated with these neurodegenerative diseases in the region and provide insights into some of the clinical presentations observed within the studied families, thereby enhancing our understanding of AD and FTD across this vast and diverse population. By expanding the genomic dataset in the coming years, we aim to deepen our understanding of the genetic underpinnings and clinical presentations of these neurodegenerative diseases across diverse Latin American populations, enhancing the potential for targeted interventions and therapies.

## Methods

### A. Participant recruitment

Participants with mild to moderate Alzheimer’s disease (AD) or frontotemporal dementia (FTD) were recruited from ten urban memory clinics across six Latin American countries: Argentina, Brazil, Chile, Colombia, Mexico, and Peru. Recruitment occurred in two phases due to the COVID-19 pandemic, which delayed the start of ReDLat’s prospective recruitment, originally planned for 2020. Despite the timing differences, the inclusion criteria for both the retrospective and prospective cohorts were identical.

All participants underwent medical and neuropsychological evaluation. The clinical diagnosis was determined by site investigators through consensus conferences at each site, adhering to the current diagnostic criteria for AD and FTD. (16–18) Healthy controls were recruited at the same locations meeting the following criteria: Clinical Dementia Rating (CDR) (19) of 0, a Mini-Mental State Examination (MMSE) (20) score greater than 25, or having been evaluated by a neuropsychologist who confirmed normal cognition in participants with few years of formal education. Family members of participants with AD or FTD, aged 18 years or older, were included if there were two or more individuals with neurodegenerative illnesses in the family, or were related to a study participant with a known dementia-associated genetic mutation and having undergone genetic counseling. All participants (diagnosed patients, healthy controls, and family members) demonstrated minimum fluency in the language of assessment (Spanish or Portuguese), had adequate vision and hearing for cognitive testing as determined by the investigator, and were required to have a study partner (informant) with at least six months of knowledge about their daily activities and cognitive/functional status. All participants (prospective and retrospective) had to be capable of providing informed consent or have a legally authorized representative. Consent was obtained following a detailed explanation of the procedures, associated risks, and potential benefits. This process complied with the ethical guidelines of each participating country and the International Declaration of Helsinki, including securing assent from the participants themselves, ensuring they expressed their willingness to participate. The study and informed consent were approved by the Institutional Review Board of each participating site.

### B. Clinical characterization

Recruited participants underwent a family history in which information was collected from patients and their study partners via self-report in the Genetic Pedigree Software-Progeny®. (21) A positive family history was defined as having at least one first- or second-degree relative with dementia or another neurodegenerative disorder. Families with three or more affected individuals in two consecutive generations were then labeled as ‘strong family aggregation’. Medical history and a full neuropsychological examination were conducted as described in Ibañez et. al. (2021). (14) Retrospective participants were assessed based on a re-evaluation of the available clinical data for each individual. The cognitive tests were harmonized as described in Maito et.al.(2023). (22)

### C. Genetic sequencing and data processing

1. Sample acquisition and processing: Standardized phlebotomy with EDTA tubes was used for sample collection. DNA was extracted using the techniques detailed in the Supplementary table 1. Samples were shipped quarterly from the various participating sites in Latin America to the United States. HudsonAlpha Institute for Biotechnology (Alabama, U.S.A) performed Single Nucleotide Polymorphism (SNP) Arrays, whole exome sequencing (WES), and/or whole genome sequencing (WGS) of the samples. Additional whole genome sequencing was performed at Psomagen, Inc. (Maryland, U.S.A)
2. SNP Arrays: Variants were genotyped using the NeuroBooster array from Illumina, designed to to capture variants relevant to neurological conditions. (23) Quality control (QC) of the SNP Array data was conducted using Genotools v1 default settings. (24) Prior to imputation, the QC’ed output files from Genotools were processed with the *no_qc_imputation_prep.sh* script. This script ran the datasets through the Wrayner script to compare them against all TOPMed freeze 8 variants. Excluded variants were then flipped to rescue additional variants, after which the dataset was processed again through the Wrayner script to compare against PASS TOPMed freeze 8 variants. Data was subsequently imputed using the TOPMed Imputation Panel and Server v1.3.3, following a previously developed pipeline for multi ancestral sample sets as described in Vitale et. al. (2024). (24)
3. WES: DNA was processed using Integrated DNA Technologies xGen Exome Hyb Panel v2, and sequenced on the NovaSeq 6000 platform using paired-end 100-base pair reads to a target depth of 100X.
4. WGS: Samples at Psomagen were prepared using the TruSeq DNA PCR-Free library prep method to avoid PCR amplification bias. Samples at HudsonAlpha underwent a custom PCR-free preparation involving Covaris shearing (fragmenting the DNA), end repair (preparing the DNA fragments for ligation), and adapter ligation, all without PCR amplification. All libraries from both sites were then normalized using KAPA qPCR and sequenced on the Illumina NovaSeq 6000 platform to a target depth of 30X. The sequencing was paired-end with a read length of 150 bp (Illumina 150bpPE).
5. Alignment and variant calling: The raw sequence data (fastq files) were aligned to the hg38 reference genome using the Sentieon v202112.05 implementation of the BWA MEM algorithm at HudsonAlpha. Sentieon v202112.05 utilities were used to sort the reads, mark duplicate sequences, and recalibrate the base quality scores. Variant calling, which identifies differences between the sample DNA and the reference genome, was performed using GATK4 tools implemented by Sentieon v202112.05. This step was conducted across all samples in a single batch to maintain consistency. Finally, variant quality score recalibration (VQSR) was applied to filter out false positive variant calls, ensuring high-quality data. This comprehensive approach achieved an average recall rate of 99.22% when compared to the Genome in a Bottle high confidence truth sets, indicating a high level of accuracy in detecting genetic variants.

### D. Genomic analyses

1. Genomic data quality control:

a. Variant Call Format (VCF) files were filtered according to established criteria to ensure high-quality data. For whole genome and exome sequences variants with genotype quality greater than 20 and read depth scores above 10 were retained. The filtered VCF was then annotated with gene names, variant types, and amino acid changes for all exonic variants using GRCh38.99 with SnpEff, dbSNP release 156, CADD 1.6, TOPMed Bravo Freeze 8 allele frequencies, and ClinVar through bcftools. (25–28) Variants with genotyping rates below 95% by individual and 95% by variant were removed. Chromosomal sex was further validated via genetic data by splitting the pseudoautosomal regions of the X chromosome and analyzing the heterozygosity of X-chromosome, as well as the count of variants present on the Y chromosome. A detailed pipeline and scripts are available at https://github.com/TauConsortium/redlat-genetics
b. Relatedness: Family history was documented through elaboration of detailed pedigrees for each recruited participant. Disclosed relatedness was compared to expected genetic relatedness using KING. (29) Individuals with cryptic relatedness (kinship coefficient <0.125 without a familial relationship documented on the pedigrees) or discrepancies between disclosed and genetic relatedness were removed.
c. Combining Datasets: To combine the arrays and WGS data, all variant IDs in both datasets were first annotated with “chrom:pos:ref:alt”. The VCFs were then intersected using BCFtools v1.9 isec, producing a list of intersecting variants between the two datasets. Both VCFs were filtered to include only these intersecting variants and then merged using BCFtools v1.9 merge. (30) After merging, the VCF was annotated with dbSNP 156. (31)
d. Concordance Check: To ensure concordance between the imputed arrays and WGS, concordance was checked for individuals with data from both methods. The imputed array data was filtered for varying allele frequencies (AFs) using BCFtools v1.9. (30) Concordance was assessed for each AF-filtered VCF using SnpSift concordance. The final concordance for each individual was determined by dividing the number of correct variant calls by the total number of intersecting variant calls. The complete script for the concordance check is available at the project’s GitHub repository [see data sharing]. Samples with concordance below 0.95 at sites with AFs less than 0.0001 were excluded.
2. Population Stratification:

a. Principal Component Analysis (PCA) was conducted using the smartpca package from EIGENSOFT (version 8.0.0). (32) Initial PCA calculations were based on samples from the 1000 Genomes Project, (33) and the ReDLat samples were subsequently projected onto the top principal components.
b. Global ancestry was estimated using ADMIXTURE (version 1.3). (34) Initially, we performed an unsupervised ancestry analysis on data from the 1000 Genomes Project, which included individuals from various ‘reference populations’. (33) The ReDLat samples were then projected onto the reference panel-based population structure.
3. Variant pathogenicity analysis: Data from WGS and WES was merged for a joint analysis for pathogenic variation. We used the Online Mendelian In Men (OMIM) database to search for genes associated with autosomal dominant, autosomal recessive, or X-linked forms of adult-onset dementia. (35) We manually curated protein-altering variants in the ten genes most commonly associated with adult-onset neurodegeneration: *APP*, *CHMP2B*, *FUS*, *GRN*, *MAPT*, *PSEN1*, *PSEN2*, *TARDBP*, *TBK1*, and *VCP*, as well as expansions in *C9orf72*. (36) Variants located in introns, the 3’ untranslated region (3’ UTR), the 5’ untranslated region (5’ UTR), and synonymous variants within exons were included if their *in silico* splice-predicting scores (dbscSNV_RF_SCORE and dbscSNV_ADA_SCORE) were both greater than 0.6, since these variants were considered likely to have an impact on splicing, making them relevant for further study. (37) Exonic non-synonymous variants (missense, nonsense, and frameshift) were analyzed following guidelines from the American College of Medical Genetics and Genomics (ACMG) [20] and the Guerreiro algorithm for *PSEN1* and *PSEN2* genes. [21] The variants identified in the remaining genes listed in <u>Supplementary Table 2</u> were queried in ClinVar [18] and AlzForum [19] and reported if previously classified as pathogenic or likely pathogenic. Variants identified through this process were then tested by hand for familial segregation to confirm their association with the disease within families.

## Results

### A. Characterization of the population

By the time of manuscript submission, we had sequenced a total of 2,254 participants from both the retrospective and prospective cohorts. Following thorough quality control, genomic data from 2,162 individuals were retained for analyses. The final dataset comprised 658 participants with SNP array data (including 174 who also had WES), 1,495 with WGS, and 9 individuals with only WES. [<u>Figure 1</u>] Among the participants with high quality genomic data, 999 were diagnosed with AD, 381 with FTD, and 755 were classified as healthy at the time of evaluation. Additionally, the sample included eight participants with mild cognitive impairment and 19 with conditions attributed to other neuropsychiatric factors (including Parkinson’s disease, Lewy body dementia, atypical parkinsonism, neurodevelopmental disorders, cerebellar ataxia, brain tumor, cognitive impairment associated with non-brain cancer, vascular dementia, severe depressive disorder, bipolar disorder, obstructive sleep apnea, and chronic traumatic encephalopathy). These individuals were part of the recruited families and had a recruited relative with AD or FTD related dementia.

**Figure 1.**
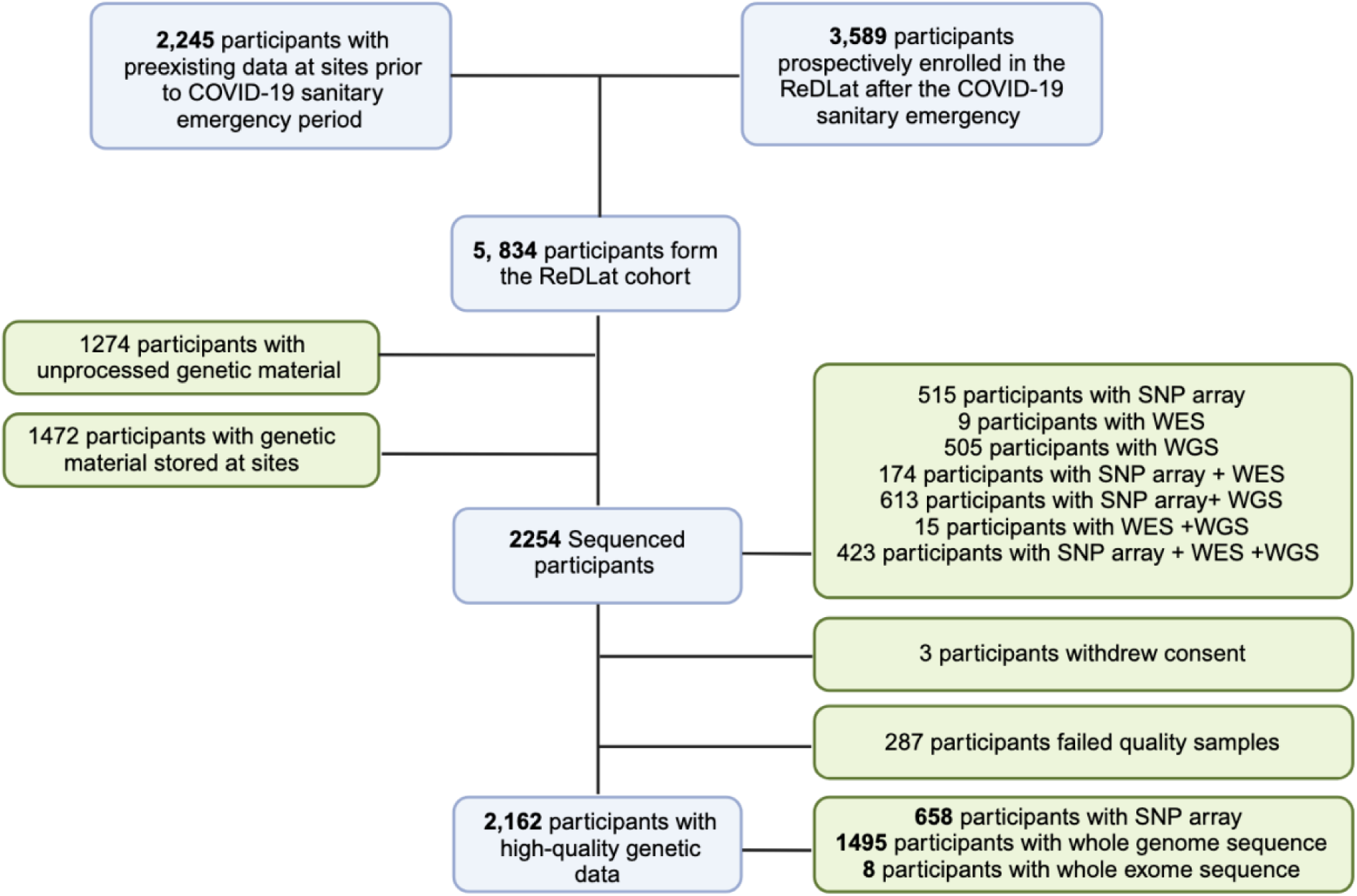
Consort Diagram of the ReDLat Cohort. **SNP:** Single Nucleotide Polymorphism, **WES:** Whole exome sequencing, **WGS:** Whole genome sequencing

As expected, a higher percentage of participants with FTD were under 65 years of age compared to those with AD (22.3% vs. 13.9%). Furthermore, the percentage of female participants was higher in the AD group (66.6%) compared to the FTD group (52.8%). Those in the AD group also showed a higher proportion of individuals who are heterozygous (41.5%) and homozygous (8.6%) for the *APOE*ℇ4 allele. [<u>Table 1</u>] The distribution of *APOE* alleles of unrelated individuals varies per country as shown in supplementary table 3. Homozygous *APOEℇ2* carriers were observed only in Colombia (0.2%) and Brazil (1%). Conversely, the highest numbers of *APOEℇ4* alleles were found in Argentina and Colombia (39% in both), although Colombia has 1% more homozygous carriers than Argentina. It is worth noting that these numbers may not be fully representative due to the Colombian sample size being considerably larger. As larger numbers of samples from all regions are collected, allele frequency estimates will be further refined.

**Table 1.**
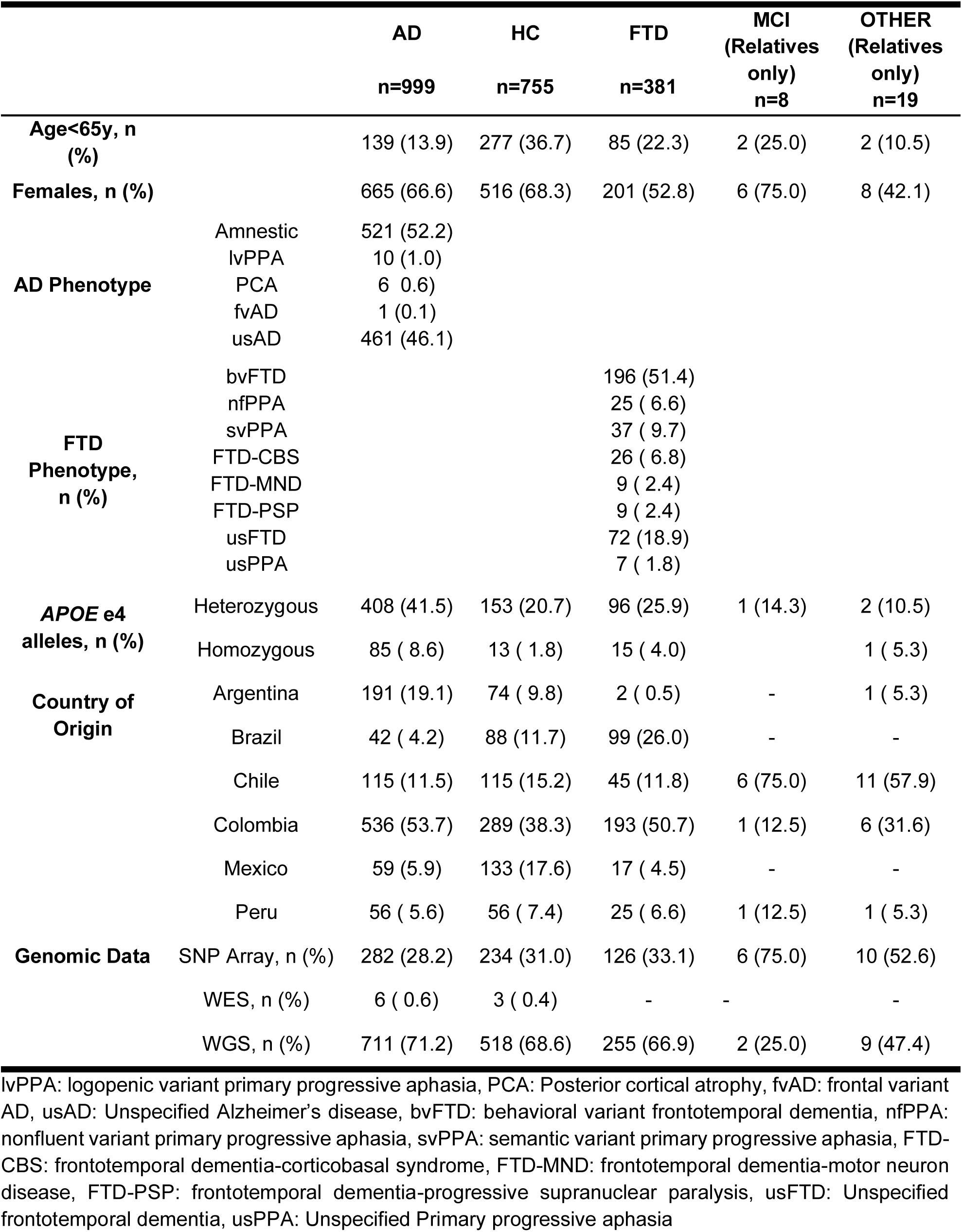
Clinical characteristics of all included ReDLat participants.

|  |  | AD | HC | FTD | MCI<br>(Relatives<br>only)<br>n=8 | OTHER<br>(Relatives<br>only)<br>n=19 |
| --- | --- | --- | --- | --- | --- | --- |
|  |  | n=999 | n=755 | n=381 |  |  |
| **Age<65y, n (%)** |  | 139 (13.9) | 277 (36.7) | 85 (22.3) | 2 (25.0) | 2 (10.5) |
| **Females, n (%)** |  | 665 (66.6) | 516 (68.3) | 201 (52.8) | 6 (75.0) | 8 (42.1) |
| **AD Phenotype** | Amnestic | 521 (52.2) |  |  |  |  |
|  | lvPPA | 10 (1.0) |  |  |  |  |
|  | PCA | 6 (0.6) |  |  |  |  |
|  | fvAD | 1 (0.1) |  |  |  |  |
|  | usAD | 461 (46.1) |  |  |  |  |
| **FTD Phenotype, n (%)** | bvFTD |  |  | 196 (51.4) |  |  |
|  | nvPPA |  |  | 25 (6.6) |  |  |
|  | svPPA |  |  | 37 (9.7) |  |  |
|  | FTD-CBS |  |  | 26 (6.8) |  |  |
|  | FTD-MND |  |  | 9 (2.4) |  |  |
|  | FTD-PSP |  |  | 9 (2.4) |  |  |
|  | usFTD |  |  | 72 (18.9) |  |  |
|  | usPPA |  |  | 7 (1.8) |  |  |
| **APOE e4 alleles, n (%)** | Heterozygous | 408 (41.5) | 153 (20.7) | 96 (25.9) | 1 (14.3) | 2 (10.5) |
|  | Homozygous | 85 (8.6) | 13 (1.8) | 15 (4.0) |  | 1 (5.3) |
| **Country of Origin** | Argentina | 191 (19.1) | 74 (9.8) | 2 (0.5) | - | 1 (5.3) |
|  | Brazil | 42 (4.2) | 88 (11.7) | 99 (26.0) | - | - |
|  | Chile | 115 (11.5) | 115 (15.2) | 45 (11.8) | 6 (75.0) | 11 (57.9) |
|  | Colombia | 536 (53.7) | 289 (38.3) | 193 (50.7) | 1 (12.5) | 6 (31.6) |
|  | Mexico | 59 (5.9) | 133 (17.6) | 17 (4.5) | - | - |
|  | Peru | 56 (5.6) | 56 (7.4) | 25 (6.6) | 1 (12.5) | 1 (5.3) |
| **Genomic Data** | SNP Array, n (%) | 282 (28.2) | 234 (31.0) | 126 (33.1) | 6 (75.0) | 10 (52.6) |
|  | WES, n (%) | 6 (0.6) | 3 (0.4) | - | - | - |
|  | WGS, n (%) | 711 (71.2) | 518 (68.6) | 255 (66.9) | 2 (25.0) | 9 (47.4) |

lvPPA: logopenic variant primary progressive aphasia, PCA: Posterior cortical atrophy, fvAD: frontal variant AD, usAD: Unspecified Alzheimer's disease, bvFTD: behavioral variant frontotemporal dementia, nvPPA: nonfluent variant primary progressive aphasia, svPPA: semantic variant primary progressive aphasia, FTD-CBS: frontotemporal dementia-corticobasal syndrome, FTD-MND: frontotemporal dementia-motor neuron disease, FTD-PSP: frontotemporal dementia-progressive supranuclear paralysis, usFTD: Unspecified frontotemporal dementia, usPPA: Unspecified Primary progressive aphasia

### B. Genetic Ancestry

To assess genetic ancestry similarity among our samples, we initially generated a merged ReDLat dataset that included 2,153 participants with WGS or SNP array data that passed the concordance analysis [Supplementary file 1]. We then used WGS from the 1000 Genomes Project (1000GP) [16] as reference populations to estimate the global ancestry of the participants, employing Principal Component Analysis (PCA) and ADMIXTURE software to estimate global ancestry. (34) We used the 1000GP cohort to identify variants with allelic frequency >10% and in linkage equilibrium that were present in both the 1000GP and the merged ReDLat dataset, resulting in a total of 226,524 variants used for ancestry analyses.

PCA [<u>Figure 2</u>, and <u>Supplementary Figure 1</u>] reveals that the ReDLat dataset shows substantial overlap with the American populations (AMR) sampled by the 1000GP, which also included Colombian, Mexican, and Peruvian participants. Though a substantial number of participants overlap with European (EUR) cohorts, there were 21 individuals clustering with the East Asian (EAS) population, suggesting recent EAS descent subgroups within ReDLat.

**Figure 2.**
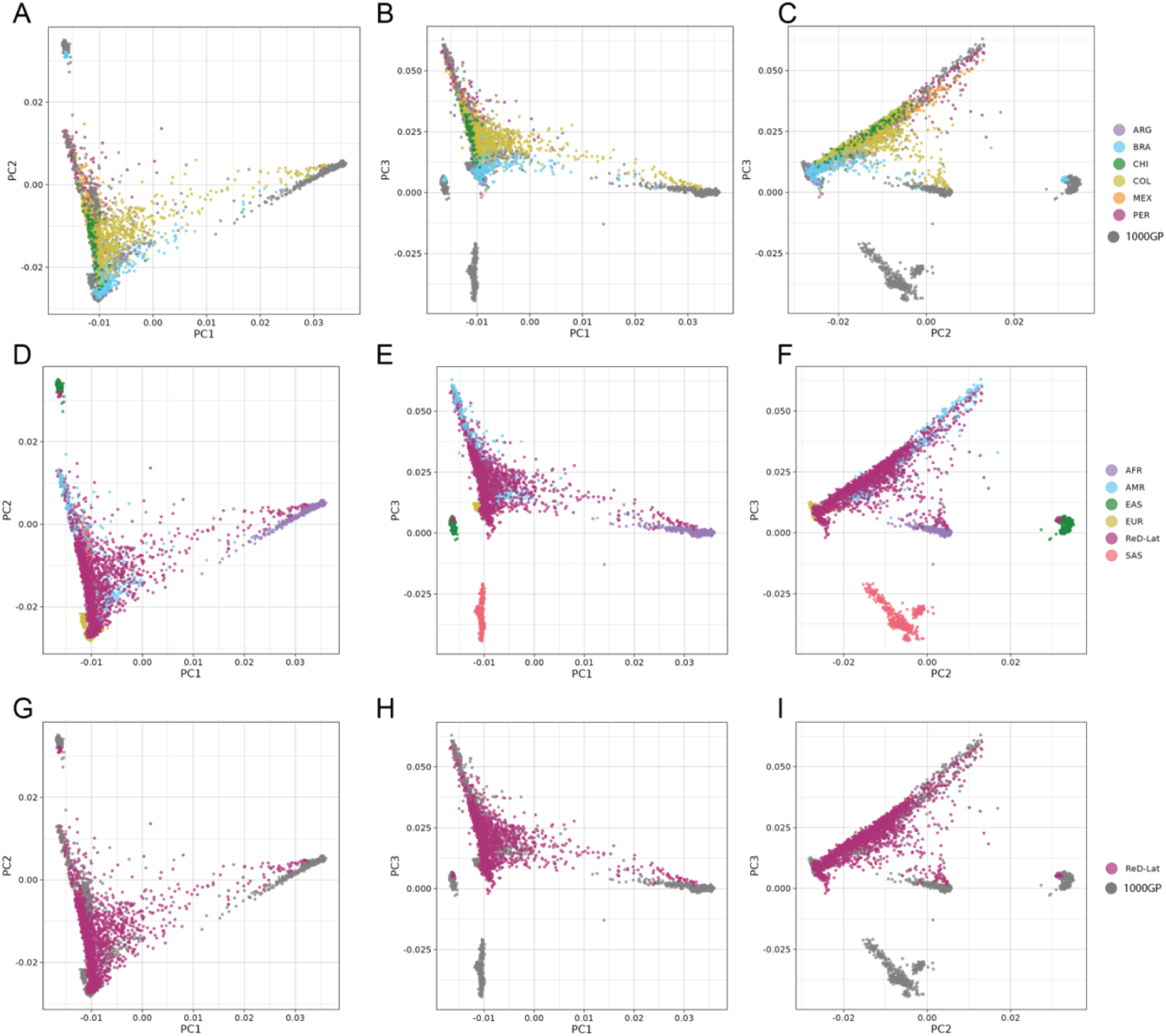
Principal component analysis (PCA) of the ReDLat cohort. **A-B-C:** ReDLat sub-cohort distribution compared to the 1000GP. **D-E-F:** 1000GP sub-cohort distribution compared to ReDLat. **G-H-I:** ReDLat compared to the 1000GP. **A-D-G:** PC1 vs. PC2. **B-E-H:** PC1 vs. PC3. **C-F-G:** PC2 vs. PC3 1000GP: Reference dataset from the 1000 Genomes project; AFR: African, AMR: American admixed, EAS: Eastern Asia, EUR: Europe, SAS: South Asian. ReDLat subcohorts include ARG: Argentina, BRA: Brazil, CHI: Chile, COL: Colombia, MEX: Mexico, PER: Peru

When analyzing the data by country, participants from Peru, Mexico, Chile, and Argentina exhibit minimal variation along Principal Component 1 (PC1), which is associated with African ancestry. In contrast, there is greater variation along Principal Component 2 (PC2), which is associated with Amerindian ancestry. This variation is particularly pronounced among participants from Mexico and Peru, who have individuals with a majority of their ancestry being Amerindian. These findings suggest a predominant two-way ancestry pattern in these populations. There is clear overlap between Argentinian and Brazilian samples with the European (EUR) cohort; however, the Brazilian samples show significant variation along PC1, highlighting the African component of the sample. While most Brazilian samples are distributed primarily between African (AFR) and EUR populations, a small subset clusters with EAS, indicating a distinct ancestral subgroup within Brazil. Colombia displays a clear three-way admixture pattern, as evidenced by its wide distribution along both PC1 and PC2. Additionally, Peruvian and Mexican samples from ReDLat exhibit clear overlap with their 1000GP counterparts, while ReDLat Colombian samples showed greater diversity than those in 1000GP [<u>Figure 2</u>]. This increased diversity is likely due to ReDLat’s broader sampling across multiple regions of Colombia. Overall, the PCA analysis confirms that the ReDLat cohort accurately represents the different historical admixture patterns previously described in the corresponding countries. (1)

To calculate global ancestry Q-values, which are adjusted p-values accounting for multiple testing and controlling the false discovery rate, we projected the ReDLat samples onto the 1000GP dataset ADMIXTURE results at multiple clustering values. <u>[Supplementary Figure 2]</u>. At K=5, where K represents the number of ancestral populations in the clustering analysis, we observed a continental separation of ancestral origins and were able to differentiate the Amerindian component. <u>[Figure 3]</u> Peru is the only country where Amerindian ancestry exceeds European ancestry, followed by Mexico, where these two ancestries show similar distributions. In Argentina, Brazil, Colombia, and Chile, European ancestry is the most prevalent among the participants, with a median value of 86.9% (Mean value of 79.7%, Standard deviation 17.5) for Argentina and 84.2% (Mean value of 73.8%, Standard deviation 26.9) for Brazil. African ancestry is present in Colombia and Brazil at lower levels; we observe a continuum of this ancestry, with individuals having over 90% and 75% African descent, down to the mean levels for both countries (around 10%), suggesting ongoing admixture over generations. In contrast, this continuum is not observed in the EAS component of the Brazilian samples, suggesting a recent diaspora without intercontinental admixture. <u>[Supplementary Figure 3]</u>

**Figure 3.**
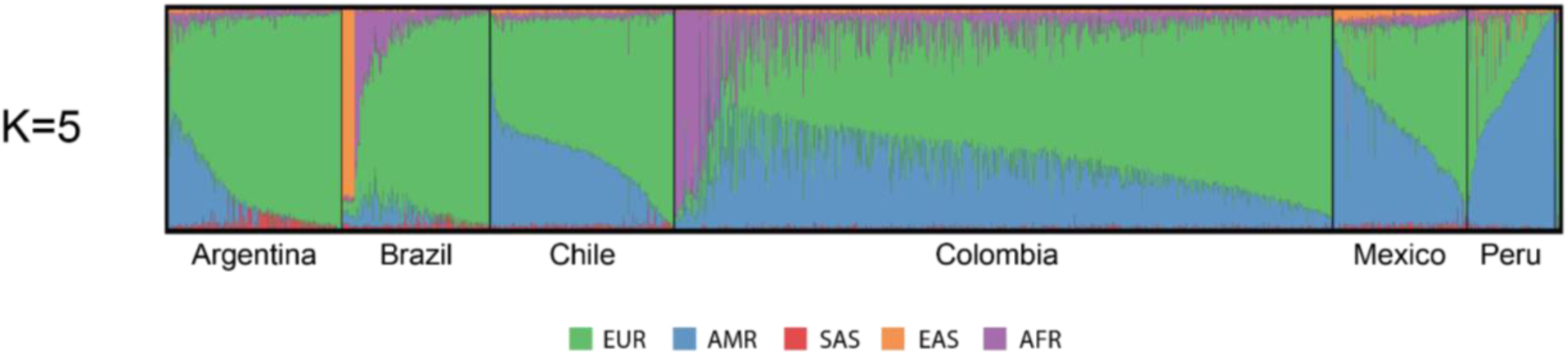
Global ancestry of ReDLat cohorts. Reference panel from the 1000 Genomes project; AFR: African, AMR: American admixed, EAS: Eastern Asia, EUR: Europe, SAS: South Asian. (see supplementary figure2)

### C. Variant pathogenicity analysis

To identify Mendelian forms of neurodegeneration in our cohort, we analyzed data from 1,678 participants who had high-quality WGS or WES data to detect pathogenic variants. Following standard practices in complex systems analysis, we employed both “bottom-up” and “top-down” approaches.

Our bottom-up approach was a "gene-to-family" search, in which we initially assessed genes most commonly associated with adult-onset neurodegeneration for pathogenic variants (see Methods). We identified a total of 17 pathogenic variants, a pathogenic *C9orf72* expansion, and 44 variants of uncertain significance (VUS). [<u>Table 2</u> and <u>Supplementary Table 4</u>]. In families with AD, we discovered a novel variant in the *PSEN1* gene, c.519G>T(p.Leu173Phe), as well as other previously described variants in the region, such as c.356C>T (p.Thr119Ile). (5) While variants like c.428T>C (p.Ile143Thr) and c.356C>T (p.Thr119Ile) are identical by descent due to founder effects, another variant, c.415A>G (p.Met139Val), has been identified as identical by state across multiple ancestral origins and presents as either AD or atypical dementia [<u>Clinical Vignette 1</u>] In families with FTD, the gene most commonly associated with hereditary forms of the illness in our cohort was *GRN*, followed by *MAPT* [<u>Clinical Vignette 2</u> and <u>Clinical Vignette 3</u>]. Pathogenic *C9orf72* repeat expansions were observed in several families from geographically non-adjacent areas.

**Table 2.**
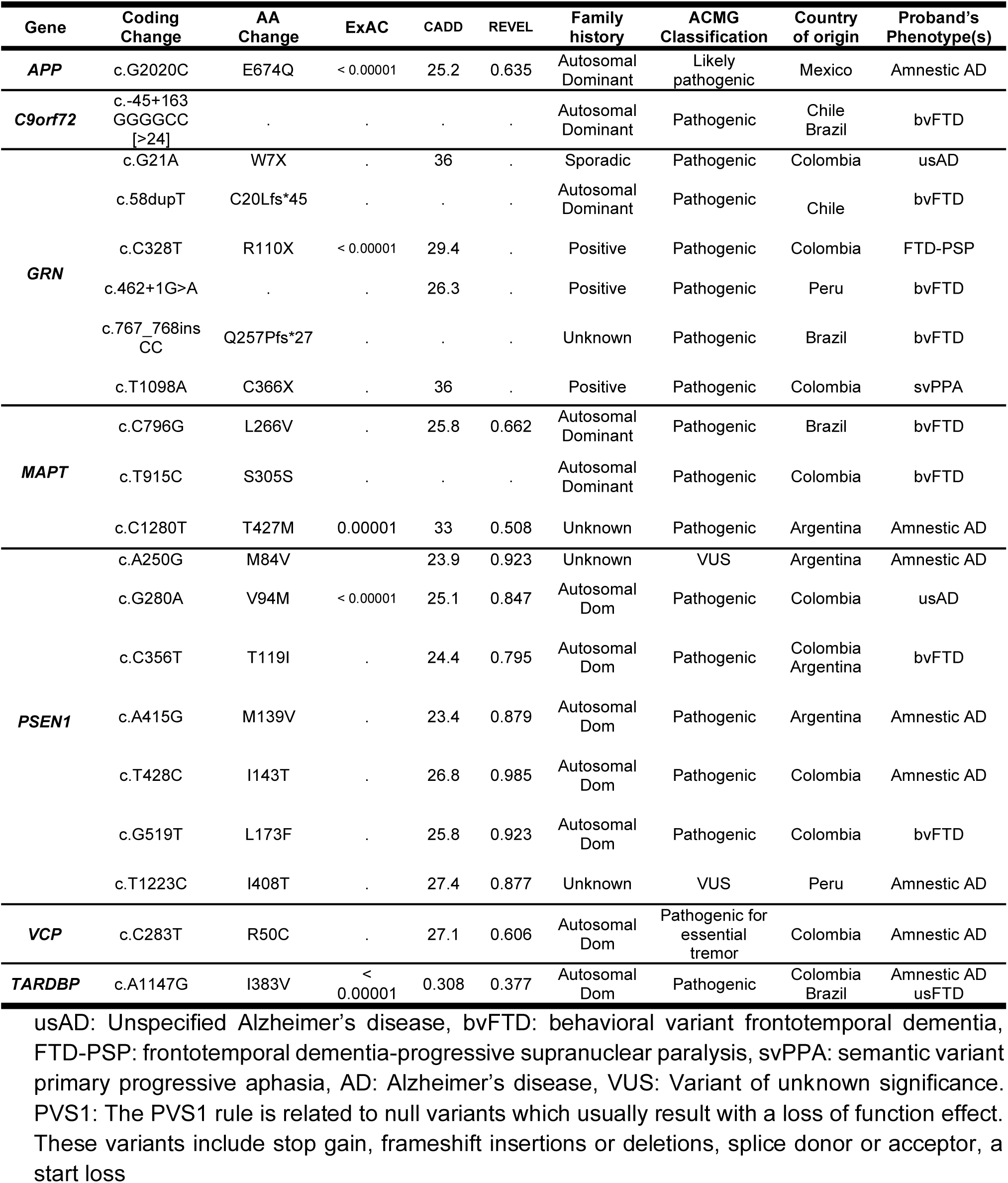
Pathogenic variants found in primary AD/FTD genes.

After the initial analysis of these primary genes, we expanded our search to include secondary genes associated with adult-onset neurodegeneration. Utilizing the OMIM database, we identified a set of genes where single nucleotide variants or short insertions/deletions could be disease-causing [<u>Supplementary Table 2</u>]. (5,38–42) In our analysis, we found four additional pathogenic variants.<u>[Supplementary table 5]</u> Three of these variants are present in families with autosomal dominant disease and are located in the *PRNP* and *NOTCH3* genes. Most notably, we identified an FTD patient without motor symptoms carrying a pathogenic variant in *SOD1* c.388G>A (p.Phe21Leu) which was previously reported in another FTD patient from the same geographical region <u>[Supplementary Table 6]</u>. (5)

In contrast, our top-to-bottom approach consisted of a “family-to-gene” search, where we analyzed the pedigrees of all recruited participants. After excluding individuals recruited as “healthy”, we identified 426 independent families that included a relative with neurodegeneration, who was considered the proband, and classified them based on the presence of affected relatives [<u>Figure 4</u>]. Among our cohort, 70 families exhibited autosomal dominant inheritance of neurodegenerative diseases, as evidenced by the presence of three affected individuals in two consecutive generations. The families were later classified according to the diagnosis of the proband, with 48 families identified as having AD and 22 as FTD. These families were subsequently categorized based on the age at disease onset: ‘late onset’ was assigned to families where all affected members presented dementia at ages older than 65 years; ‘early onset’ applied to those where all affected individuals were 65 years or younger at the time of dementia onset; and ‘mixed onset’ described families that included members with both early and late-onset disease <u>[Supplementary Table 7]</u>. Though many of the carriers of pathogenic variants belonged to the RedLat retrospective cohort, 14 of the recruited families were carriers of pathogenic variants, and the majority had positive family history of neurodegeneration.

**Figure 4.**
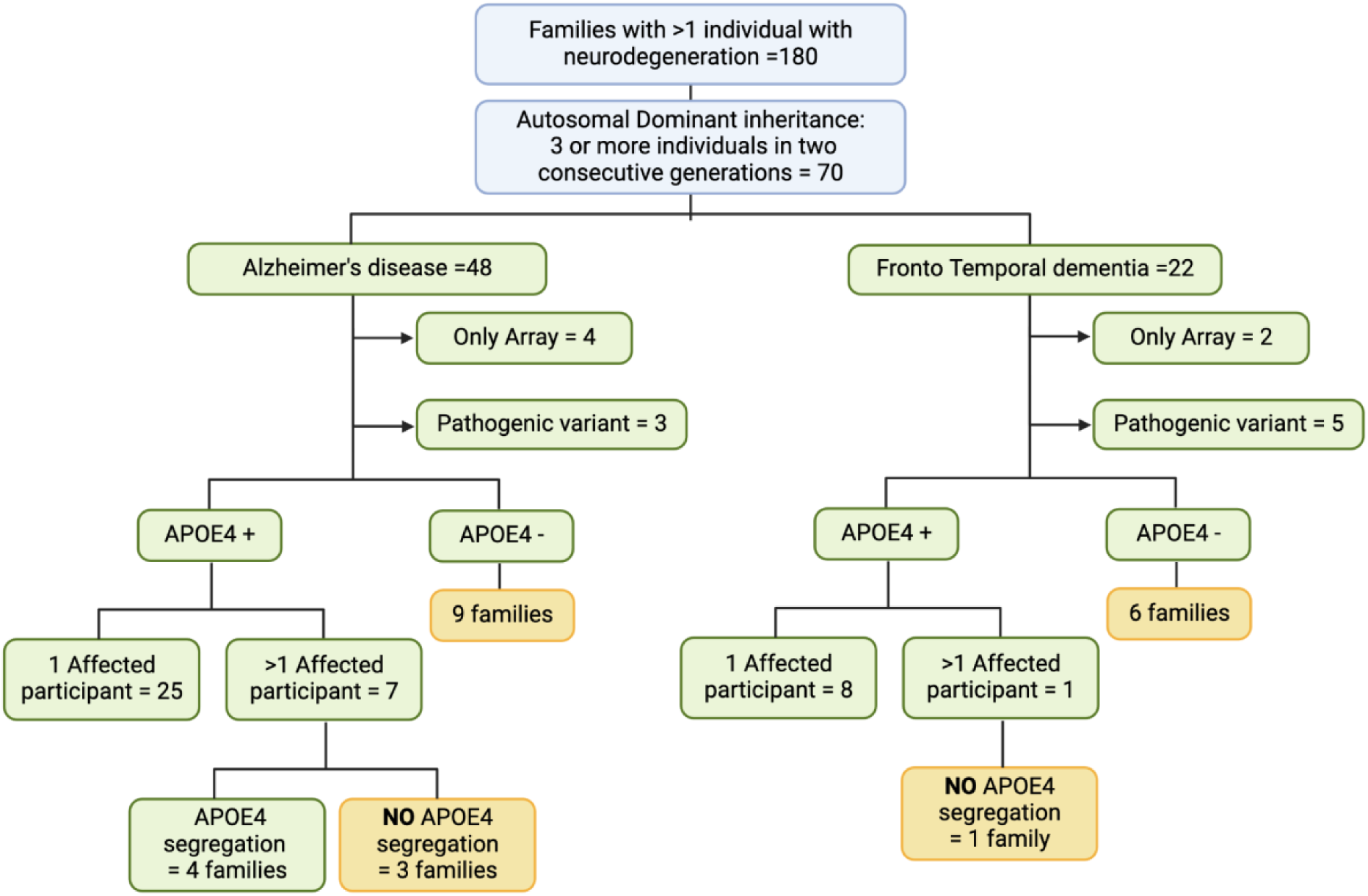
Segregation of neurodegeneration in the ReDLat recruited cohort.

In a further analysis of families exhibiting autosomal dominant patterns, we assessed the presence of at least one allele of *APOE ℇ4*. Among the families diagnosed with AD, nine were negative for *APOE ℇ4* alleles, while 32 had ar least an *APOE ℇ4* carrier. Among these families we determined the *APOE ℇ4* allele status for more than one participant in seven of them. Notably, only four families demonstrated a segregation pattern consistent with the presence of the *APOE ℇ4* allele in the affected individuals. In contrast, among the families with FTD, six were negative for *APOE ℇ4* alleles, and the one family where we could determine the *APOE ℇ4* allele status for more than one participant, no segregation pattern was observed. <u>[Figure 4]</u>

## Discussion

This initial release of genomic data from the ReDLat cohort provides early insights into the genetic underpinnings of neurodegeneration within a Latin American population, supported by genomic analyses of established variants associated with AD and FTD. Our genetic ancestry analysis, leveraging data from the 1000 Genomes Project, revealed tricontinental admixture patterns across most regions and an East Asian component in Brazil, reflecting historical migration and admixture events. The study notably identifies a significant prevalence of autosomal dominant inheritance patterns in AD and FTD, characterized by distinct age-of-onset categorizations, geographic distribution of genetic variants, and with a stronger presence of the *APOE*ℇ4 allele in AD families. These patterns also include newly discovered variants in the *PSEN1* and *APP* genes for AD, which play critical roles in the disease’s pathogenesis. Families with as-yet unidentified variation remain strong candidates for future novel gene discovery as additional family members are recruited for gene-mapping linkage studies.

Indeed, there is considerable potential for novel genetic discovery in diverse cohorts such as ReDLat, both in terms of risk for ADRD and resilience against it, in both families and sporadic cases. As demonstrated by the *PSEN1* E280A kindred from Colombia,(43) leveraging larger, diverse cohorts–as well as genetic families with substantial clinical heterogeneity–represents a unique opportunity for discovery of resilience factors for ADRD, which may serve as strong targets for disease intervention.

As previously noted by Browning et. al. (2018), historical factors like colonization, migration, and genetic bottlenecks have significantly shaped the genetic landscape of Latin American populations. During and after the period of colonization, many Latin Americans lived in small, often isolated villages. This created a population structure characterized by multiple mini-bottlenecks, as descendants of a small number of founders in each village tended to remain in the same location for many generations. As families expanded, the specific rare alleles in each place became fixed and more observable. This phenomenon resulted in a genetic map of the region that closely corresponds to the geographic map, facilitating genetic discovery. The long stretches of identical genes (long runs of homozygosity) and increased diversity within isolated populations are advantageous for researchers seeking to identify rare variants associated with diseases like AD and FTD. These factors underscore the value of family studies in Latin America, offering unique insights into genetic patterns and the potential for discovering new genetic contributions to disease. (44)

The first-wave study cohort reported here has several limitations. First, we have chosen not to conduct unbiased discovery efforts, such as genome-wide association studies and burden analyses, in this cohort due to the extensive family structure and relatively small sample size of the cross-sectional cohort collected to date. Second, despite the tangible advancement in global representation offered by this cohort, participants are still enriched for higher socioeconomic status due to the urban-centric recruitment. This drawback is being actively addressed through ongoing enrollments and community outreach efforts in more rural areas. Third, as with any clinic-based enrollment cohort, there is a possibility of ascertainment bias among recruited participants because the study recruits from clinical practices specializing in cognitive disorders, which may lead to an overrepresentation of more extreme clinical phenotypes.

## Conclusion

We present here the first snapshot of genetic contributors to dementias from a distributed cohort across Latin America. The findings to date represent significant advances in understanding the etiology of Alzheimer’s and Frontotemporal dementia in this region. Continued enrollment in this project will provide additional valuable insights through future studies that map the genetic underpinnings of disease risk in large families, genetic risk burden in cases, and offer well-powered cohorts for case-control studies to identify common risk variants. Moreover, the robust family structure already observed in ReDLat provides a unique opportunity to map genetic modifiers and assess the impact of local genomic ancestry on these modifiers. As global population representation continues to expand, it will be critical to evaluate the generalizability of genetic risk factors for AD and FTD across diverse ancestral backgrounds, within the context of distinct social determinants of health, and accounting for modifiable risk factors that may influence disease risk and resilience across distinct cultures.

## Data Availability

All the scripts used for the study have been compiled into a single GitHub repository: https://github.com/TauConsortium/redlat-genetics. Data from the present study is available upon request to the authors and the executive committee of the Multi-Partner Consortium to Expand Dementia Research in Latin America (ReDLat)

## List of abbreviations

*1000GP:*: 1000 Genomes Project
*ACE-III:*: Addenbrooke’s Cognitive Examination-III
*ACMG:*: American College of Medical Genetics
*AD:*: Alzheimer’s disease
*AFR:*: African
*AF:*: Allelic frequency
*CDR:*: Clinical Dementia Rating Scale
*DNA:*: Deoxyribonucleic acid
*EDTA:*: Ethylenediaminetetraacetic acid
*EUR:*: European
*FTD:*: Frontotemporal dementia
*GIAB:*: Genome in a Bottle Consortium
*MoCA:*: Montreal Cognitive Assessment
*MMSE:*: Mini-Mental State Examination
*OMIM:*: Online Mendelian Inheritance in Men database
*PC:*: Principal component
*PCA:*: Principal component analysis
*PCR:*: Polymerase chain reaction
*QC:*: Quality control
*qPCA:*: Quantitative Polymerase chain reaction
*ReDLat:*: Multi-Partner Consortium to Expand Dementia Research in Latin America
*SNP:*: Single Nucleotide Polymorphism
*SNV:*: Single nucleotide variants
*VCF:*: Variant Call Format
*VUS:*: Variant of uncertain significance
*VQSR:*: Variant quality score recalibration
*WES:*: Whole exome sequencing
*WGS:*: Whole genome sequencing

## Declarations

### A. Ethics approval and consent to participate

Written informed consent following the guidelines of the Code of Ethics of the World Medical Association, Helsinki Declaration and Belmont Report was obtained from all participants or their legally authorized proxies for all evaluations and assessments conducted. This consent included explicit permission to publish the findings.The project was approved by the Institutional Review Board of Each Medical Institution:

1. Argentina: INECO-Centro de Psicología Médica San Martín de Tours: FWA00028264
2. Brazil: Hospital das Clínicas da Faculdade de Medicina da Universidade de São Paulo: FWA00001035
3. Chile:Hospital Clínico Universidad de Chile: FWA00029089
4. Chile:Universidad Adolfo Ibanez. FWA00030846
5. Colombia: Comité de Bioética del Instituto de Investigaciones Médicas, Facultad de Medicina, Universidad de Antioquia: FWA00028864
6. Colombia: Pontificia Universidad Javeriana - Hospital Universitario San Ignacio: FWA00001113
7. Colombia: Fundación Valle de Lili: FWA00029865
8. Mexico: Instituto Nacional de Ciencias Médicas FWAA00014416
9. Peru: Hospital Nacional Docente Madre Niño San Bartolome FWA00010121
10. USA: University of California San Francisco-Memory and Aging Center: FWA00000068

### B. Consent for publication

Not applicable

### C. Availability of data and materials

All the scripts used for the study have been compiled into a single GitHub repository: https://github.com/TauConsortium/redlat-genetics

### F. Authors’ contributions

J.A-U, S.D.P-E., J.S.Y and K.S.K. conceived and designed the project with some discussion from J.N.C., J.W.T, A.P.C., C.J., D.L.M., and M.L.N. E.A.B. handled the samples, and K.R. generated the SNP-array data at HudsonAlpha. All authors read and approved the final manuscript.

## Acknowledgements

We thank the individuals and the families who participated in this study.

## G. Authors’ information

Juliana Acosta-Uribe and Stefanie D. Piña-Escudero contributed equally to this work Jennifer S. Yokoyama and Kenneth S. Kosik contributed equally to this work

## H. Footnotes

Not applicable

## Clinical Vignettes

### Clinical Vignette 1

#### Family with *PSEN1* c.415A>G (p.Met139Val)

This vignette describes a family with early-onset Alzheimer’s disease (EOAD), where the disease presented similarly across all affected members. Initial symptoms appeared between ages 35 and 45 and included temporal and spatial disorientation, speech disturbances, and memory difficulties. Early evaluations revealed declines in social cognition, phonological and semantic verbal fluency, and memory. As the disease progressed, there were notable reductions in processing speed, phonological verbal fluency, visuoconstructive abilities, and working memory. In advanced stages, severe impairments in memory, task-switching, processing speed, and phonological fluency severely impacted communication and daily functioning. Additional neurological symptoms included vascular episodes, hallucinations, violent behavior, spasmodic movements, and seizures.

Genomic analysis identified a pathogenic variant in *PSEN1* c.4.415A>G (p.Met139Val), which was present in more than 10 family members, confirming a genetic basis for the disease.

The MRI of one of the Met139Val carriers showed cortical atrophy, cavum septi pellucidi (arrow head), and hippocampal volume reduction (arrow).

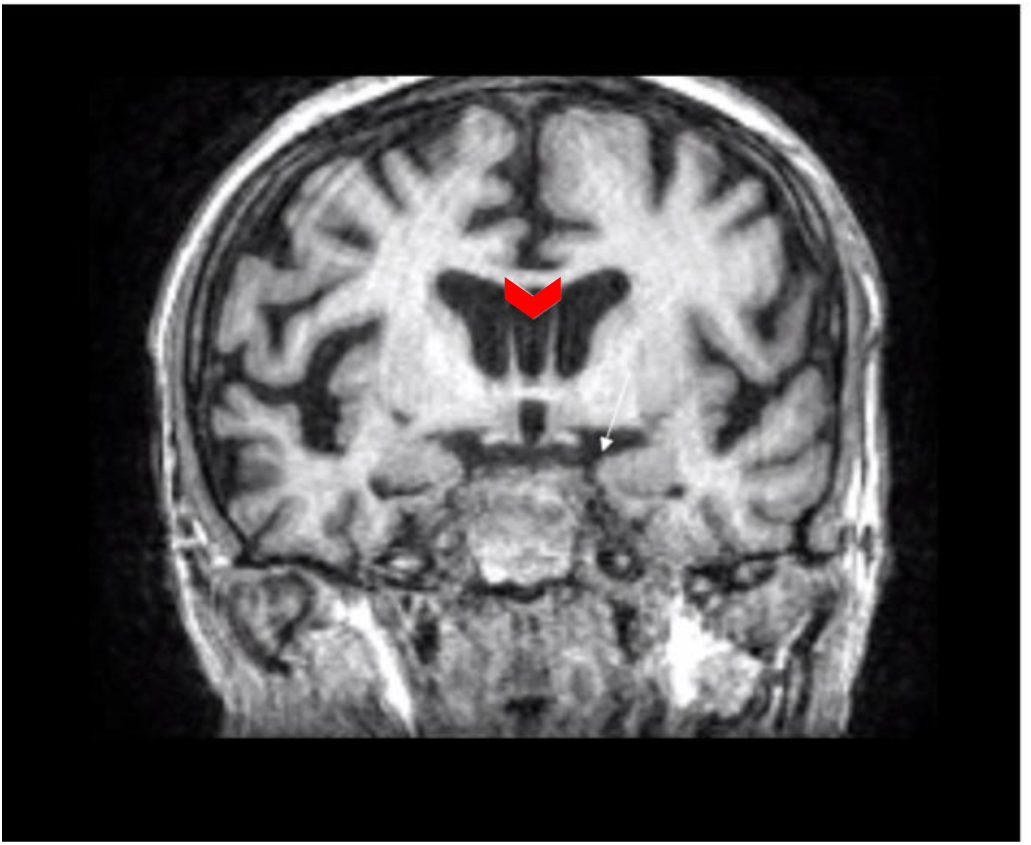

An anatomopathological study of this same person later revealed a positive immunohistochemistry for β-amyloid and phosphorylated tau (p-tau) in the hippocampus and cortex, indicating significant neurodegenerative changes confirming Alzheimer’s disease as a result of the genetic variant in this family.

### Clinical vignette 2

#### Clinical presentation of a family with *MAPT* c.915T>C: (p.Ser305Ser)

This vignette describes a family with more than ten individuals affected by Frontotemporal dementia (FTD), presenting either as behavioral variant FTD (bvFTD) or as progressive supranuclear palsy (PSP). The proband, a 45–50-year-old individual, had a family history marked by severe behavioral changes in multiple relatives. Their first symptom, problem-solving difficulty around age 45-50, was a common initial sign among family members. As the disease progressed, they experienced loss of figurative language comprehension and gait changes. Shared symptoms across affected family members included fixations on specific foods, hyperphagia, and increased consumption of sweets. Social cognition impairments were marked by difficulties in social interactions, inappropriate behaviors, reduced emotional responses to others’ pain, and notable aggressiveness. Behaviorally, individuals engaged in repetitive activities, neglected personal appearance, and showed hoarding tendencies. A unique symptom across all affected individuals was a persistent habit of tapping their teeth until they loosened and could be pulled out.

### Clinical Vignette 3

#### Clinical presentation of a family with *MAPT* c.796C>G:(p.Leu266Val)

The proband presented to the clinic with a five-year history of progressively worsening behavioral changes. Initial symptoms were difficult to assess, partly due to their living situation with an adolescent son who could not provide reliable information. The patient had become increasingly reliant on Post-it notes to remember tasks and was easily irritable. Their living conditions had severely deteriorated, with moldy, larval-infested clothing and significant neglect of household responsibilities, ultimately leading them to move in with a sibling by the age of 30-35.

During an initial assessment in early 2019, the patient required assistance with dressing and bathing and had started using diapers. Apathy was the predominant neuropsychiatric symptom, with occasional agitation, though they were generally quiet and withdrawn. Their appetite fluctuated between periods of anorexia and hyperphagia, during which they consumed inappropriate substances, such as skincare products and soap.

Trazodone was prescribed for insomnia, which also helped manage episodes of aggression. The patient had a long-standing history of alcoholism and had been involved in a car accident around age 25-30 due to this issue. The disease progressed rapidly, with the MMSE score dropping from 13 to untestable within six months. By this stage, the patient exhibited speech stereotypy, frequently repeating phrases like ‘banana with milk’ and ‘I want to see my son today.

Blood tests revealed a positive antinuclear antibody test (>1/1280), positive anti-thyroid antibodies, and mildly elevated ammonia levels, with no other signs of autoimmune or hepatic conditions detected. MRI scan indicated severe frontotemporal atrophy (arrows), predominantly affecting the anteroinferior and medial parts of the temporal lobes(arrow heads) predominately on the right side.

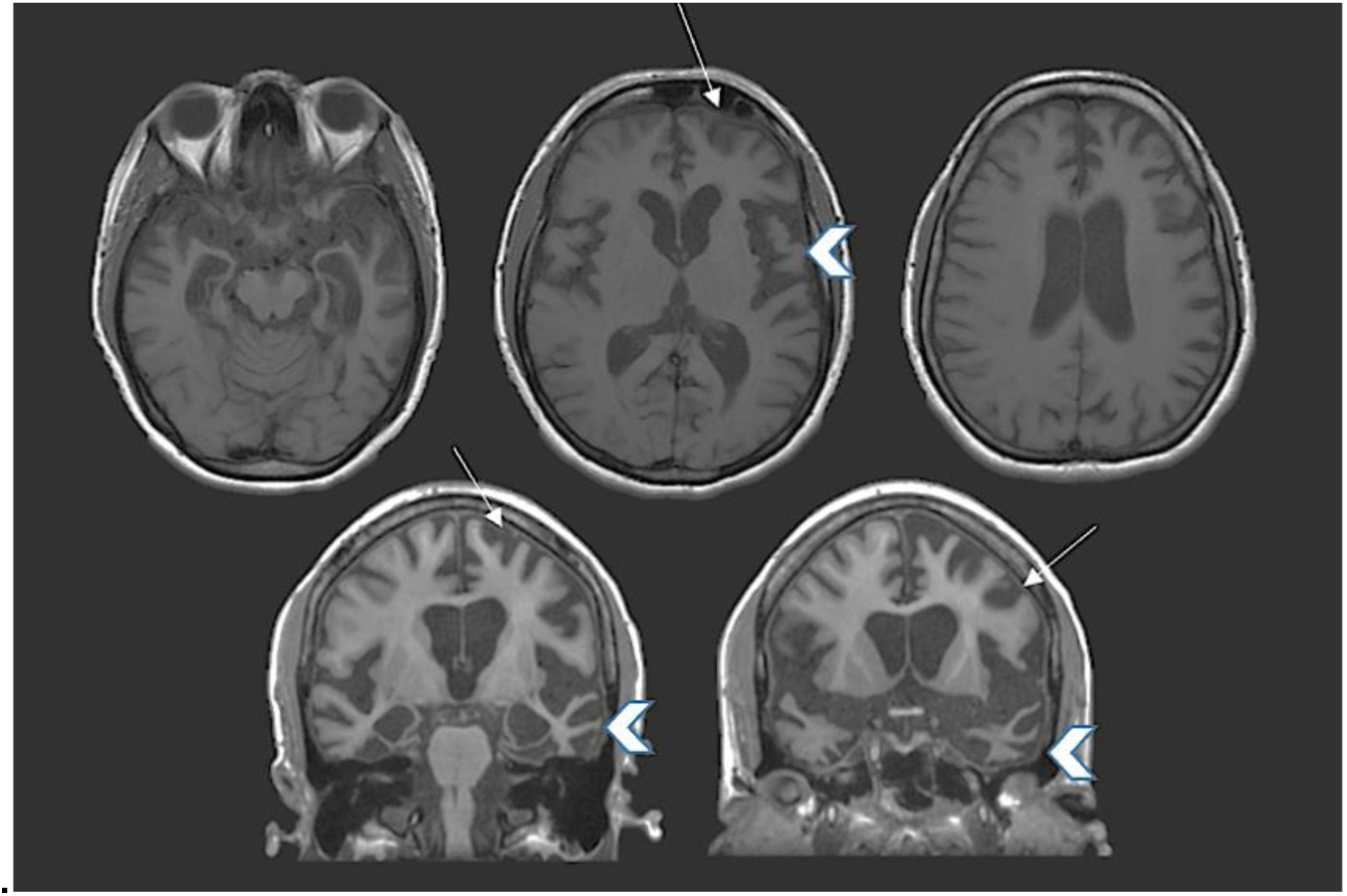

The participant was identified as heterozygous for a pathogenic variant in the *MAPT* gene, specifically c.796C>G (p.Leu266Val). The patient passed away at the age of 30-35 as a result of multiple organ failure secondary to a COVID-19 infection.

The proband’s parent passed away before the age of 40-45, initially diagnosed with schizophrenic catatonia, though their symptoms were more consistent with behavioral variant frontotemporal dementia (bvFTD). Despite having young children, the parent would leave the house at 6 am, returning only at 10 pm. The family later discovered that they had been begging for money from strangers, smoking discarded cigarettes, and consuming food found on the ground. Additionally, the participant’s grandparent and two relatives also died before the age of 40-45, exhibiting psychiatric symptoms or signs of early-onset dementia.

## Supplementary Tables

**Supplementary Table 1.** DNA extraction techniques per site.

| Site | DNA extraction kit | Extraction Protocol |
| --- | --- | --- |
| **Colombia** | The *Wizard*® Genomic DNA Purification Kit | Manufacturer recommendations |
| Bogotá |  |  |
| Medellín |  |  |
| **Peru** |  |  |
| **Chile** |  |  |
| Santiago East |  |  |
| Santiago West |  |  |
| **Brazil** |  |  |
| **Colombia** | QIAamp® DNA Mini Kit | Manufacturer recommendations |
| Cali |  |  |
| **Argentina** |  |  |
| Buenos Aires |  |  |
| San Juan |  |  |
| **Mexico** | in-house developed DNA extraction technique | Starting Volume: Add a total of 100 uL of molecular grade water (H<sub>2</sub>O).<br>Sample Preparation: Dilute up to 50 mL with blood without plasma using Lysis Buffer (prepared with Tris base 8, sucrose, magnesium chloride, and Triton X-100). Mix for 10 minutes and centrifuge for 10 minutes at 3500 rpm at room temperature.<br>First Centrifugation: Decant the supernatant and adjust the remaining volume to 25 mL with Lysis Buffer.<br>Resuspension: Vortex until the pellet is dispersed and centrifuge again for 10 minutes at 3500 rpm at room temperature.<br>Second Centrifugation: Carefully decant the supernatant without disturbing the pellet and allow the contents to dry.<br>Digestion Setup: In an Eppendorf tube, add 5 microliters of Proteinase K (PK), 160 microliters of PK buffer, 80 microliters of 10% SDS, and 490 microliters of molecular biology grade water.<br>Sample Transfer and Incubation: Transfer the pellet to the Eppendorf tube containing the previous substances. Vortex and incubate for 2 hours at 55°C.<br>Salt Precipitation: Add 400 microliters of 3M NaCl and vortex. Centrifuge at the maximum speed of a microcentrifuge (15,000 rpm) for 10 minutes.<br>Ethanol Precipitation: Transfer the contents to a cytometry tube with 1800 microliters of absolute ethanol. Mix gently and observe the DNA strand.<br>DNA Pelleting: Centrifuge at 3500 rpm for 15 minutes and discard the supernatant.<br>Ethanol Wash: Add 300 microliters of 70% ethanol. Centrifuge at 3500 rpm for 5 minutes and discard the supernatant.<br>Drying: Allow drying for 10 minutes.<br>Rehydration: Add 100 uL of H<sub>2</sub>O. Incubate at 37°C for 2 hours.<br>Final Transfer: Transfer the contents to an Eppendorf tube.<br>DNA Concentration Measurement: Measure the concentration of DNA. |

**Supplementary table 2.** Genes associated with adult-onset dementia and/or neurodegeneration.

| Gene | Location | Phenotype | Inheritance | Phenotype MIM |
| --- | --- | --- | --- | --- |
| *AARS1* | 16q22.1 | Leukoencephalopathy, hereditary diffuse, with spheroids | AD | 619661 |
| *ALS2* | 2q33.1 | Amyotrophic lateral sclerosis, juvenile | AR | 205100 |
| *ANG* | 14q11.2 | Amyotrophic lateral sclerosis | AD | 611895 |
| *ANXA11* | 10q22.3 | Amyotrophic lateral sclerosis | AD | 617839 |
| *ATP13A2* | 1p36.13 | Kufor-Rakeb syndrome | AR | 606693 |
| *ATP7B* | 13q14.3 | Wilson disease | AR | 277900 |
| *C19orf12* | 19q12 | Neurodegeneration with brain iron accumulation | AD, AR | 614298 |
| *CCNF* | 16p13.3 | Frontotemporal dementia and/or amyotrophic lateral sclerosis | AD | 619141 |
| *CHCHD10* | 22q11.23 | Frontotemporal dementia and/or amyotrophic lateral sclerosis | AD | 615911 |
| *CHCHD2* | 7p11.2 | Parkinson disease autosomal dominant | AD | 616710 |
| *CLN3* | 16p12.1 | Ceroid lipofuscinosis, neuronal | AR | 204200 |
| *CLN5* | 13q22.3 | Ceroid lipofuscinosis, neuronal | AR | 256731 |
| *CLN6* | 15q23 | Ceroid lipofuscinosis, neuronal (Kufs type) | AR | 204300 |
| *CLN8* | 8p23.3 | Ceroid lipofuscinosis, neuronal, Northern epilepsy variant | AR | 610003 |
| *COASY* | 17q21.2 | Neurodegeneration with brain iron accumulation | AR | 615643 |
| *CRAT* | 9q34.11 | Neurodegeneration with brain iron accumulation | AR | 617917 |
| *CSF1R* | 5q32 | Leukoencephalopathy, diffuse hereditary, with spheroids | AD | 221820 |
| *CTSD* | 11p15.5 | Ceroid lipofuscinosis, neuronal | AR | 610127 |
| *CTSF* | 11q13.2 | Ceroid lipofuscinosis, neuronal (Kufs type) | AR | 615362 |
| *CYLD* | 16q12.1 | Frontotemporal dementia and/or amyotrophic lateral sclerosis | AD | 619132 |
| *CYP27A1* | 2q35 | Cerebrotendinous xanthomatosis | AR | 213700 |
| *DNAJC5* | 20q13.33 | Ceroid lipofuscinosis, neuronal (Kufs type), autosomal dominant | AD | 162350 |
| *DNAJC6* | 1p31.3 | Parkinson disease, early-onset | AR | 615528 |
| *EIF2B1* | 12q24.31 | Leukoencephalopathy with vanishing white matter with or without ovarian failure | AR | 603896 |
| *EIF2B2* | 14q24.3 | Leukoencephalopathy with vanishing white matter with or without ovarian failure | AR | 620312 |
| *EIF2B3* | 1p34.1 | Leukoencephalopathy with vanishing white matter with or without ovarian failure | AR | 620313 |
| *EIF2B4* | 2p23.3 | Leukoencephalopathy with vanishing white matter with or without ovarian failure | AR | 620314 |
| *EIF2B5* | 3q27.1 | Leukoencephalopathy with vanishing white matter with or without ovarian failure | AR | 620315 |
| *ERBB4* | 2q34 | Amyotrophic lateral sclerosis | AD | 615515 |
| *FBXO7* | 22q12.3 | Parkinson disease, autosomal recessive | AR | 260300 |
| *FIG4* | 6q21 | Amyotrophic lateral sclerosis | AD | 612577 |
| *FTH1* | 11q12.3 | Neurodegeneration with brain iron accumulation | AD | 620669 |
| *FTL* | 19q13.33 | Neurodegeneration with brain iron accumulation | AD | 606159 |
| *HNRNPA1* | 12q13.13 | Amyotrophic lateral sclerosis | AD | 615426 |
| *HNRNPA2B1* | 7p15.2 | Inclusion body myopathy with early-onset Paget disease with or without frontotemporal dementia | AD | 615422 |
| *HTRA1* | 10q26.13 | Cerebral arteriopathy, autosomal dominant/recessive, with subcortical infarcts and leukoencephalopathy | AD, AR | 616779 |
| *ITM2B* | 13q14.2 | Dementia, familial Danish | AD | 117300 |
| *JAM2* | 21q21.3 | Basal ganglia calcification, idiopathic, autosomal recessive | AR | 618824 |
| *KCTD7* | 7q11.21 | Epilepsy, progressive myoclonic, with or without intracellular inclusions | AR | 611726 |
| *MATR3* | 5q31.2 | Amyotrophic lateral sclerosis | AD | 606070 |
| *MFSD8* | 4q28.2 | Ceroid lipofuscinosis, neuronal | AR | 610951 |
| *MYORG* | 9p13.3 | Basal ganglia calcification, idiopathic, autosomal recessive | AR | 618317 |
| *NOTCH3* | 19p13.12 | Cerebral arteriopathy with subcortical infarcts and leukoencephalopathy | AD | 125310 |
| *OPTN* | 10p13 | Amyotrophic lateral sclerosis with or without frontotemporal dementia | AD, AR | 613435 |
| *PANK2* | 20p13 | Neurodegeneration with brain iron accumulation | AR | 234200 |
| *PARK7* | 1p36.23 | Parkinson disease, autosomal recessive early-onset | AR | 606324 |
| *PDGFB* | 22q13.1 | Basal ganglia calcification, idiopathic | AD | 615483 |
| *PDGFRB* | 5q32 | Basal ganglia calcification, idiopathic | AD | 615007 |
| *PFN1* | 17p13.2 | Amyotrophic lateral sclerosis | AD | 614808 |
| *PINK1* | 1p36.12 | Parkinson disease, early onset | AR | 605909 |
| *PLA2G6* | 22q13.1 | Neurodegeneration with brain iron accumulation | AR | 610217 |
| *PPT1* | 1p34.2 | Ceroid lipofuscinosis, neuronal | AR | 256730 |
| *PRKN* | 6q26 | Parkinson disease, juvenile | AR | 600116 |
| *PRNP* | 20p13 | Creutzfeldt-Jakob disease | AD | 123400 |
| *PTPA* | 9q34.11 | Parkinson disease, autosomal recessive early-onset, with impaired intellectual development | AR | 620482 |
| *REPS1* | 6q24.1 | Neurodegeneration with brain iron accumulation | AR | 617916 |
| *SETX* | 9q34.13 | Amyotrophic lateral sclerosis juvenile | AD | 602433 |
| *SIGMAR1* | 9p13.3 | Amyotrophic lateral sclerosis, juvenile | AR | 614373 |
| *SLC20A2* | 8p11.21 | Basal ganglia calcification, idiopathic, | AD | 213600 |
| *SNCA* | 4q22.1 | Parkinson disease | AD | 168601 |
| *SNCB* | 5q35.2 | Dementia, Lewy body | AD | 127750 |
| *SOD1* | 21q22.11 | Amyotrophic lateral sclerosis | AD, AR | 105400 |
| *SPG11* | 15q21.1 | Amyotrophic lateral sclerosis , juvenile | AR | 602099 |
| *SPTLC1* | 9q22.31 | Amyotrophic lateral sclerosis, juvenile | AD | 620285 |
| *SQSTM1* | 5q35.3 | Frontotemporal dementia and/or amyotrophic lateral sclerosis | AD | 616437 |
| *SYNJ1* | 21q22.11 | Parkinson disease early-onset | AR | 615530 |
| *TIA1* | 2p13.3 | Amyotrophic lateral sclerosis with or without frontotemporal dementia | AD | 619133 |
| *TPP1* | 11p15.4 | Ceroid lipofuscinosis, neuronal | AR | 204500 |
| *TREM2* | 6p21.1 | Polycystic lipomembranous osteodysplasia with sclerosing leukoencephalopathy | AR | 618193 |
| *TUBA4A* | 2q35 | Amyotrophic lateral sclerosis with or without frontotemporal dementia | AD | 616208 |
| *TYROBP* | 19q13.12 | Polycystic lipomembranous osteodysplasia with sclerosing leukoencephalopathy | AR | 221770 |
| *UBQLN2* | Xp11.21 | Amyotrophic lateral sclerosis, with or without frontotemporal dementia | XLD | 300857 |
| *VAPB* | 20q13.32 | Amyotrophic lateral sclerosis | AD | 608627 |
| *VPS13C* | 15q22.2 | Parkinson disease, autosomal recessive | AR | 616840 |
| *WDR45* | Xp11.23 | Neurodegeneration with brain iron accumulation | XLD | 300894 |
| *XPR1* | 1q25.3 | Basal ganglia calcification, idiopathic | AD | 616413 |

AD: Autosomal dominant, AR: Autosomal recessive, XLD: X linked dominant

**Supplementary Table 3.** *APOE* alleles per country.

| ***APOE***<br>**alleles** | **Argentina**<br>**(n=244)** | **Brazil**<br>**(n=229)** | **Chile**<br>**(n= 278)** | **Colombia**<br>**(n= 807)** | **Mexico**<br>**(n= 149)** | **Peru**<br>**(n=137)** |
| --- | --- | --- | --- | --- | --- | --- |
| e2/e2 | 0 ( 0.0) | 2 ( 0.9) | 0 ( 0.0) | 2 ( 0.3) | 0 ( 0.0) | 0 ( 0.0) |
| e2/e3 | 13 ( 5.5) | 14 ( 6.5) | 19 ( 7.0) | 57 ( 7.2) | 3 ( 2.1) | 1 ( 0.7) |
| e2/e4 | 2 ( 0.8) | 3 ( 1.4) | 4 ( 1.5) | 10 ( 1.3) | 1 ( 0.7) | 1 ( 0.7) |
| e3/e3 | 132 (55.5) | 150 (69.1) | 148 (54.8) | 385 (48.5) | 107 (73.3) | 91 (66.4) |
| e3/e4 | 75 (31.5) | 43 (19.8) | 89 (33.0) | 283 (35.6) | 31 (21.2) | 37 (27.0) |
| e4/e4 | 16 ( 6.7) | 5 ( 2.3) | 10 ( 3.7) | 57 (7.2) | 4 ( 2.7) | 7 ( 5.1) |

**Supplementary Table 4.** Summary of manual curation of primary variants in all 10 first-pass genes.

|  | Protein<br>Altering Variants | Pathogenic<br>Variants | Variants of<br>Unknown<br>Significance |
| --- | --- | --- | --- |
| *APP* | 14 | 0 | 1 |
| *CHMP2B* | 6 | 0 | 2 |
| *FUS* | 20 | 0 | 8 |
| *GRN* | 27 | 5 | 12 |
| *MAPT* | 44 | 3 | 2 |
| *PSEN1* | 17 | 7 | 2 |
| *PSEN2* | 20 | 0 | 1 |
| *TARDBP* | 3 | 1 | 1 |
| *TBK1* | 15 | 0 | 13 |
| *VCP* | 5 | 1 | 2 |
| **SUM** | 171 | 17 | 44 |

**Supplementary Table 5.** Summary of secondary variants associated with neurodegeneration.

Summary of secondary variants associated with neurodegeneration
|  | Total | Variants of<br>Unknown<br>Significance | Pathogenic<br>Variants |
| --- | --- | --- | --- |
| Non synonymous SNV | 377 | 166 | 3 |
| Exonic SNV splicing | 1 | 0 | 0 |
| Intronic SNV splicing | 14 | 3 | 0 |
| Start-loss | 4 | 1 | 0 |
| Stop-loss | 1 | 0 | 0 |
| Frameshift deletion | 7 | 2 | 0 |
| Frameshift insertion | 4 | 4 | 0 |
| Non frameshift deletion | 13 | 6 | 0 |
| Non frameshift insertion | 7 | 4 | 1 |
| **SUM** | 428 | 186 | 4 |

SNV: Single nucleotide variant

**Supplementary Table 6.** Variant filtering in secondary genes associated with neurodegeneration.

| Gene | All variation | Protein altering | Low MAF (<0.001) | CADD >20 | Clinvar Pathogenic |
| --- | --- | --- | --- | --- | --- |
| AARS1 | 14 | 7 | 1 | 0 | 0 |
| ANG | 5 | 3 | 2 | 1 | 0 |
| ANXA11 | 43 | 22 | 12 | 7 | 0 |
| C19orf12 | 8 | 3 | 3 | 3 | 0 |
| CCNF | 7 | 5 | 3 | 2 | 0 |
| CHCHD10 | 23 | 15 | 8 | 4 | 0 |
| CHCHD2 | 6 | 6 | 5 | 4 | 0 |
| CSF1R | 56 | 28 | 16 | 8 | 0 |
| CYLD | 3 | 0 | 0 | 0 | 0 |
| DNAJC5 | 9 | 2 | 2 | 1 | 0 |
| ERBB4 | 37 | 14 | 13 | 10 | 0 |
| FIG4 | 33 | 18 | 14 | 11 | 0 |
| FTH1 | 3 | 1 | 1 | 0 | 0 |
| FTL | 1 | 0 | 0 | 0 | 0 |
| HNRNPA1 | 11 | 3 | 2 | 2 | 0 |
| HNRNPA2B1 | 11 | 2 | 2 | 2 | 0 |
| HTRA1 | 9 | 2 | 0 | 0 | 0 |
| ITM2B | 15 | 9 | 8 | 5 | 0 |
| LRP12 | 7 | 3 | 2 | 2 | 0 |
| MATR3 | 20 | 16 | 15 | 12 | 0 |
| NOTCH3 | 122 | 72 | 56 | 33 | 1 |
| OPTN | 28 | 16 | 14 | 10 | 0 |
| PDGFB | 3 | 2 | 1 | 1 | 0 |
| PDGFRB | 22 | 10 | 2 | 1 | 0 |
| PRNP | 31 | 20 | 11 | 5 | 2 |
| SETX | 130 | 93 | 59 | 20 | 0 |
| SLC20A2 | 6 | 3 | 1 | 1 | 0 |
| SNCA | 8 | 5 | 4 | 4 | 0 |
| SOD1 | 3 | 2 | 2 | 2 | 1 |
| SPTLC1 | 3 | 3 | 2 | 1 | 0 |
| SQSTM1 | 42 | 24 | 16 | 9 | 0 |
| TIA1 | 7 | 3 | 0 | 0 | 0 |
| TUBA4A | 11 | 5 | 5 | 4 | 0 |
| UBQLN2 | 11 | 6 | 6 | 2 | 0 |
| VAPB | 6 | 3 | 0 | 0 | 0 |
| WDR45 | 1 | 1 | 0 | 0 | 0 |
| XPR1 | 7 | 1 | 1 | 0 | 0 |
| **Total** | 762 | 428 | 289 | 167 | 4 |

Warmer colors are used to highlight a higher number of variants in a given gene.

**Supplementary table 7.**
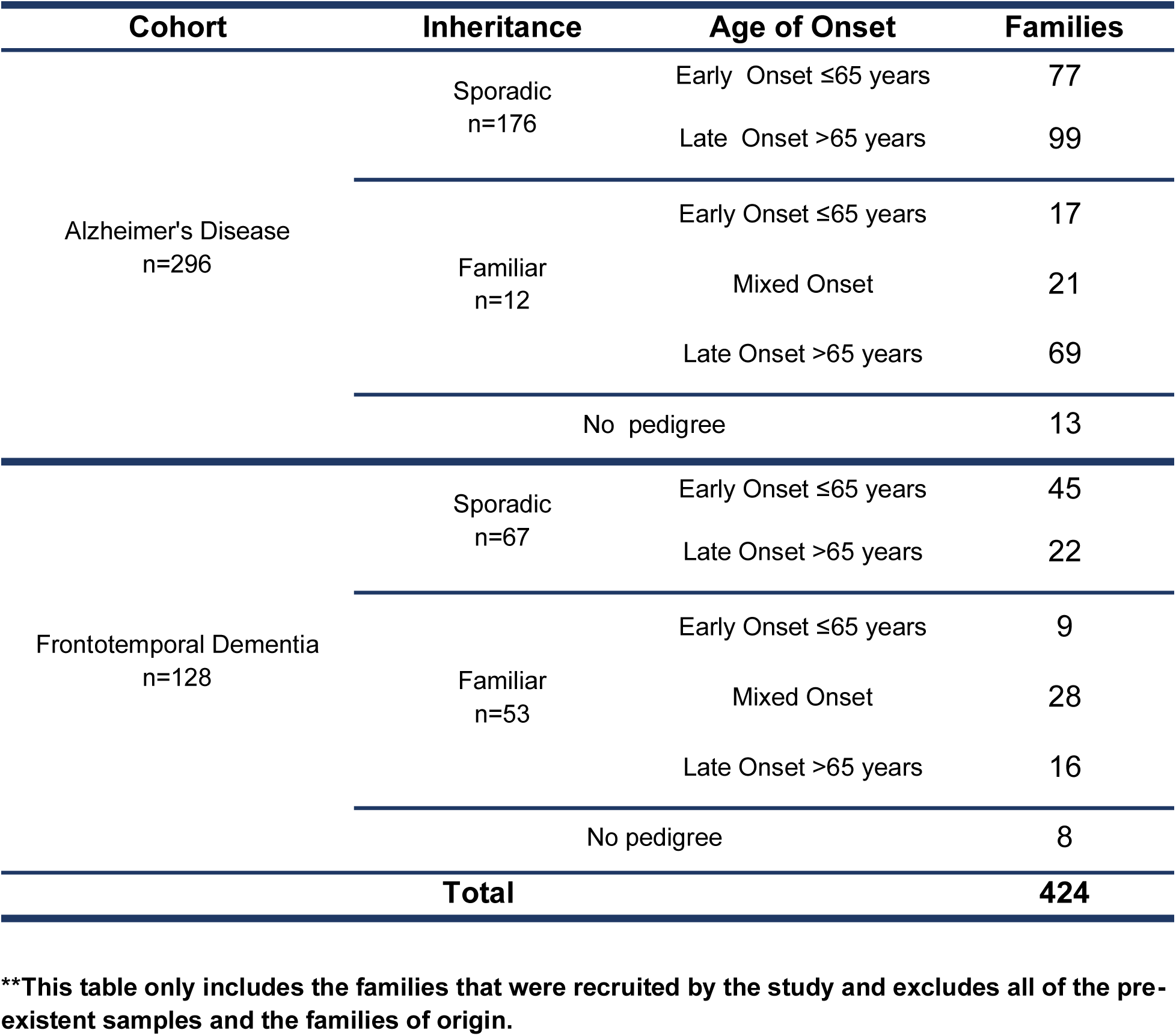
Familial clustering of Alzheimer’s disease and Frontotemporal dementia in the actively recruited participants.

## Supplementary Figures

**Supplementary figure 1.**
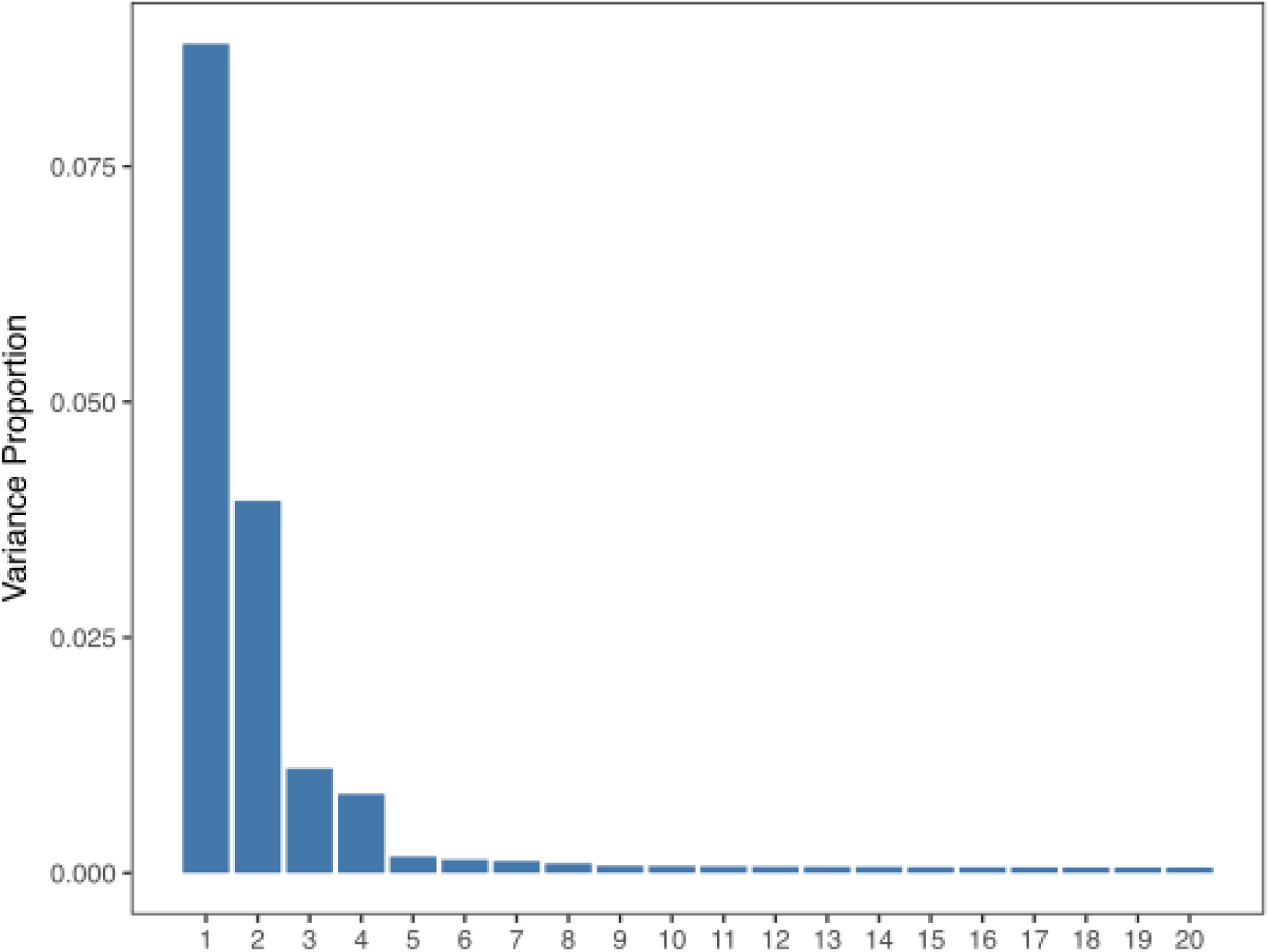
Principal component analysis scree plot. Variance explained by top 20 principal components

**Supplementary figure 2.**
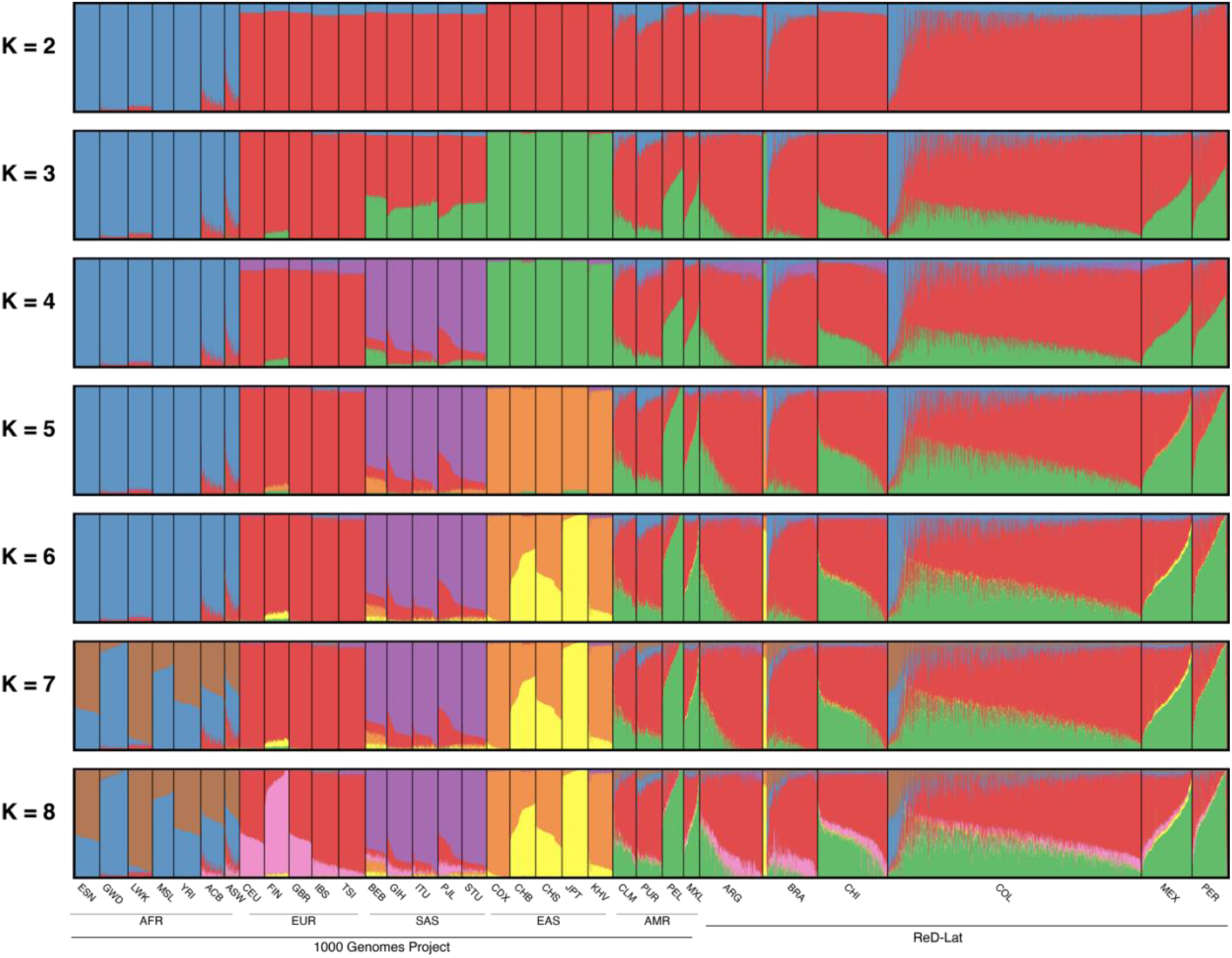
Global ancestry proportions of the ReDLat cohort represented by ADMIXTURE Q. values at different K (ancestral population) values. The 1000 Genomes project includes **African (AFR):** GWD: Gambian in Western Divisions in the Gambia, LWK: Luhya in Webuye, MSL: Mende in Sierra Leone, YRI: Yoruba in Ibadan, Nigeria, ACB: African Caribbean in Barbados, ASW: African Ancestry in Southwest US. **European (EUR):** CEU: Utah Residents (CEPH) with Northern and Western European Ancestry, FIN: Finnish in Finland, GBR: British in England and Scotland, IBS: Iberian Population in Spain, TSI: Tuscany in Italia. **South Asian (SAS):** BEB: Bengali in Bangladesh, GHI: Gujarati Indians in Houston, ITU: Indian Telugu in the U.K., PJL: Punjabi in Lahore, STU: Sri Lankan Tamil in the UK. **East Asian (EAS):** CDX: Chinese Dai in Xishuangbanna, China, CHB: Han Chinese in Beijing, CHS: Han Chinese South, JPT: Japanese, Kyushu, KHV: Kinh Vietnamese. **Admixed American (AMR):** CLM: Colombian from Medellin, PUR: Puerto Rican from Puerto Rico. **ReDLat subcohorts:** ARG: Argentina, BRA: Brazil, CHI: Chile, COL: Colombia, MEX: Mexico, PER: Peru

**Supplementary figure 3.**
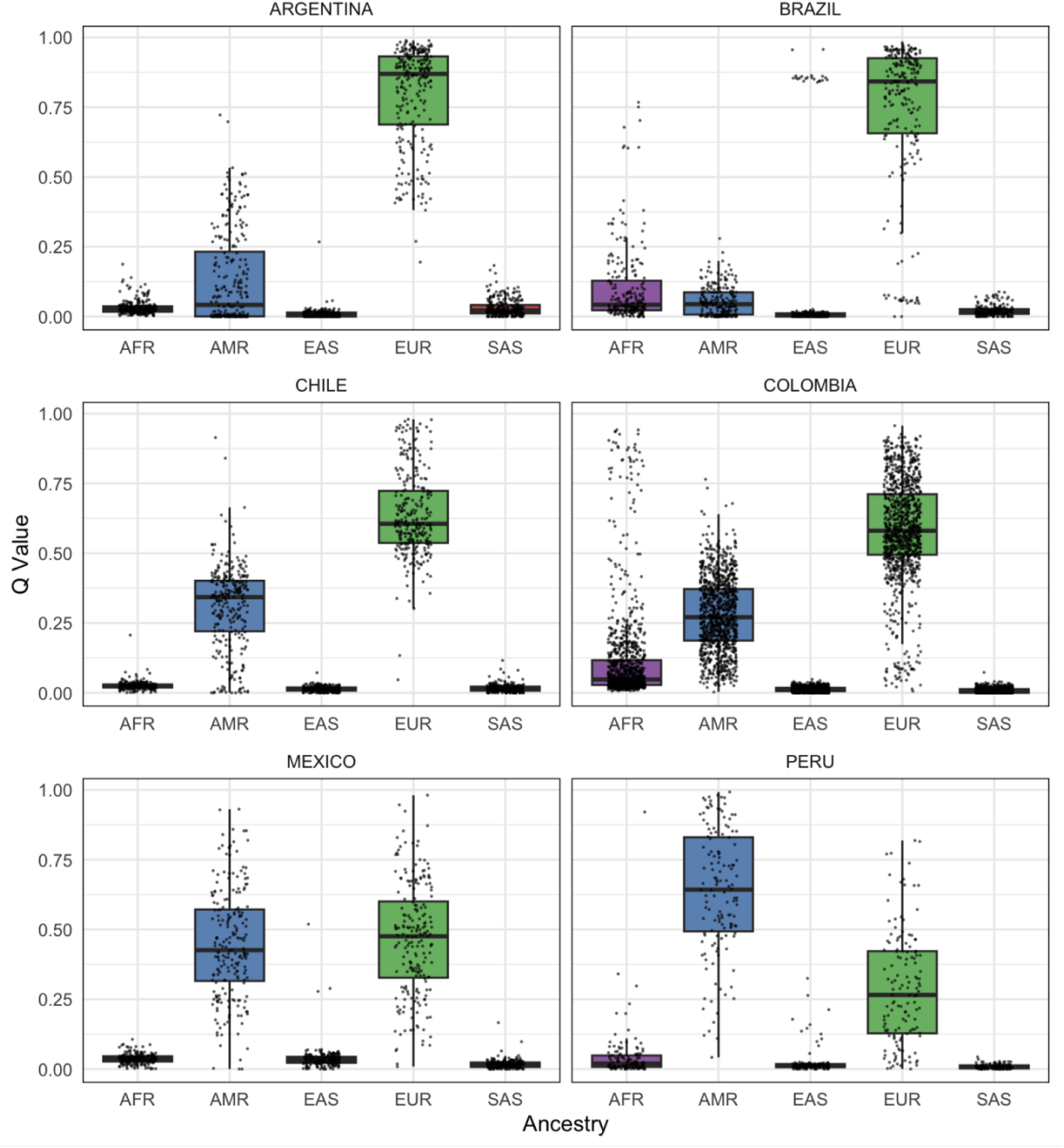
Distribution of global ancestries across the multiple countries. Calculations were based on ADMIXTURE Q. values. Reference ancestries from the 1000 Genomes project are AFR: African, AMR: American admixed, EAS: Eastern Asia, EUR: Europe, SAS: South Asian.

